# Respiratory and non-respiratory manifestations in children admitted with COVID 19 in Rio de Janeiro city, Brazil

**DOI:** 10.1101/2020.12.29.20248994

**Authors:** AR Araujo da Silva, CGB Fonseca, JLPS Miranda, BV Travassos, CR Baião, KD Silva, LBAE dos Santos, MMR de Britto, PALS Cerqueira, SNB Pereira, RBJ Rios, CS Vieira, IA Leal, NC Martins, LMAC de Carvalho, AB Pereira, CH Teixeira

## Abstract

**Introduction:** COVID 19 is still a challenge in pediatrics due to variety of symptoms and different presentations

**Aim:** To describe clinical, laboratorial and treatment of confirmed COVID-19 pediatric admitted in hospitals.

**Methods:** A retrospective study was conducted in children (0-18 years), admitted between March and November 15, 2020, with confirmed COVID-19 by reverse transcription polymerase chain reaction or serological tests. Clinical data about symptoms, laboratorial exams and treatments were analysed. Patients were evaluated according predominant (PRS) or non-predominant respiratory symptoms (non-PRS)

**Results:** Sixty-four patients were evaluated, being the median age 5.6 years. Forty-seven (73.4%) children were admitted with PRS and 17 (26.4%) with non-PRS. The main symptoms in the PRS group were fever in 74.5% of children and cough in 66%; and fever in 76.5% and edema/cavitary effusion in 29.4% in the non-PRS group. The median of C-reactive protein (in mg/dl) was 2.5 in the PRS group and 6.1 in the non-PRS group. Antibiotics were used in 85.1% of the PRS group and 94.1% of non-group. Comorbidity was present in 30/47 (63.8%) of PRS group and 8/17 (47.1%) of non-PRS group (p=0.22). Length of stay until 7 days in patients with comorbidity was present in 27/64 (42.1%) and more than 7 days in 11/64 (17.1%) (p= 0.2)

**Conclusion:** Non-PRS represented more than one quarter of admitted patients. Fever was the main symptom detected, elevated CRP was frequent and antibiotics were commonly prescribed. Comorbidity was found in both groups and his presence was not associated with a longer length of stay.

## Introduction

A cluster of pneumonia of unknown origin was described initially in late December 2019 in Wuhan, China and linked to a seafood wholesale Market ^1,2^. Just a few days later, on January 07 2020, a novel Coronavirus was identified as the causative agent and the disease experienced a rapid spread from Wuhan to the whole China in 30 days ^1,3^. The disease caused by SARS-COV-2 was named COVID 19 and during the first two months of 2020 affected at least 57 countries showing his pandemic potential ^4^. On 11th March, the World Health Organization (WHO) declared a pandemic situation due to SARS-COV-2 ^5^.

The initial reports of COVID-19 showed that disease affected more patients between 30 and 79 years, with mild cases in 81% and case fatality of 2.3% ^3^. In this same report, children and adolescents represented 2% of all patients diagnosed ^3^.

Children often have milder disease than adults and deaths have been extremely rare, although atypical manifestations as multisystem inflammatory disease, Kawasaki disease and Kawasaki-like disease have been reported ^6-9^. Newborns also have developed symptomatic COVID-19, but until the present, there is limited evidence of vertical intrauterine transmission ^10^.

Brazil is one of the most affected countries by the current pandemic. Data from Brazilian Ministry of Health reported more than 7,5 million cases and 191,570 deaths until December 28, 2020 ^11^. Despite the amount of cases, just a few reports of the behaviour of disease in Brazil were reported as case reports or children admitted to paediatric intensive care units and in pediatric hospitals ^12-15^.

Considering the relevance of SARS-COV-2 in Brazilian children, our aim is to describe the clinical, laboratorial and treatment of confirmed COVID-19 pediatric patients in Rio de Janeiro, city, Brazil.

## Methods

### Type of study

We conducted a retrospective descriptive study of pediatric patients, admitted with confirmed infection due to SARS-COV-2

### Setting

The study was done in two pediatric private hospitals from Rio de Janeiro, city. Both hospitals attend patients from 0 to 18 years old age and count with 24h-emergency service, wards, pediatric intensive care units and all pediatric specialists for support. Hospital 1 is located in the North Zone of the city and has 135 bed capacity. Hospital 2 is located in the South Zone and has 45 bed capacity. Many of the admissions are referred from peripheral hospitals from the metropolitan area of Rio de Janeiro state.

### Inclusion criteria

All patients admitted between March and November 2020, with clinical signals and symptoms suggestive of COVID-19 in the presence of positive reverse transcription polymerase chain reaction (RT-PCR) (Allplex TM, Seegene Brazil) or positive serological tests (IgM, IgG or IgA).

### Exclusion criteria

Patients admitted with clinical signals and symptoms suggestive of COVID-19 with indeterminate values of RT-PCR or serological tests or attending in the emergency department and discharged.

### Definition of clinical signals and symptoms suggestive of COVID19

The following criteria were considered as suggestive: A) Acute onset of fever AND cough OR Acute onset of ANY THREE OR MORE of the following signs or symptoms: Fever, cough, general weakness/fatigue, headache, myalgia, sore throat, coryza, dyspnoea, anorexia/nausea/vomiting, diarrhoea, altered mental status AND Epidemiological criteria. B) A patient with severe acute respiratory illness (SARI: acute respiratory infection with history of fever or measured fever of ≥ 38 C°; and cough; with onset within the last 10 days; and requires hospitalization) ^16^. Multisystem inflammatory syndrome was defined also according to WHO criteria ^17^.

### Data collection

Initially data of suspected cases were collected from Brazilian database system of electronic notification of COVID-19 (notifica.saude.gov.br). After this, all patients files were recorded in order to analyse additional signals and symptoms not included in the electronic notification as well treatments and complementary exams performed.

### Analysis of data

Patients were categorized in two groups according the main symptoms at admission: predominant respiratory symptoms and non-predominant respiratory sumptoms. Categorical variables were described as percentages and frequencies. Continuous variables were described as medians. Comparisons between groups were made using the chi-squared test or Fisher’s exact test for categorical variables, and the Mann-Whitney test for continuous variables. Shapiro-Wilk test was used to analyze the normality of the sample. A p value less than 5% was considered as statistically significant.

### Ethical aspects

The study was approved by the Ethical Committee of Federal Fluminense University, process number 4.100.232, June 20, 2020.

## Results

The first COVID19 case started the symptoms on March 25, 2020 and during the following nine months (until November 15, 2020), 74 patients were confirmed with the infection. Of all confirmed patients, 64/74 (86.5%) were admitted and 10/74 (13.5%) were referred to ambulatory follow-up and the mean age of patients (in months) was 79,4. Of 64 admitted patients, 5 (7.8%) developed symptoms of COVID19 after admission for other reasons. The median age was 67 months (5.6 years) and other demographic data of patients are shown in table 1.

**Table 1.**
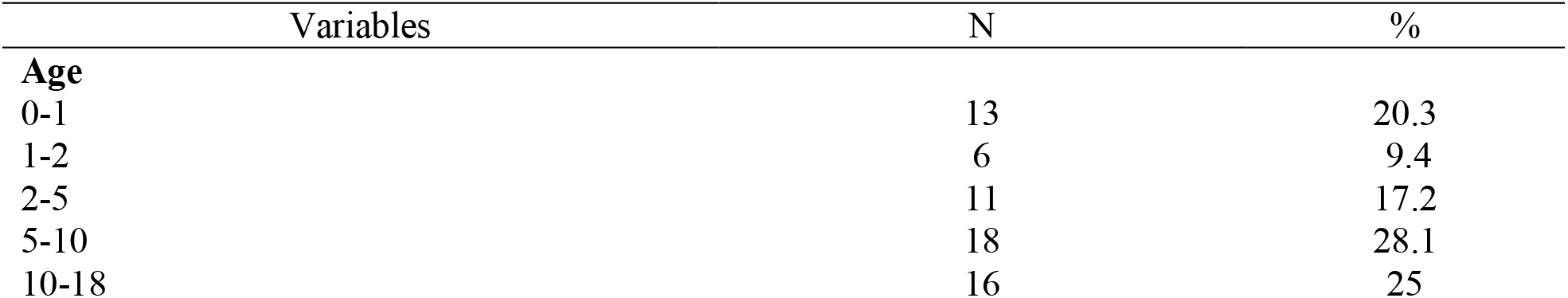
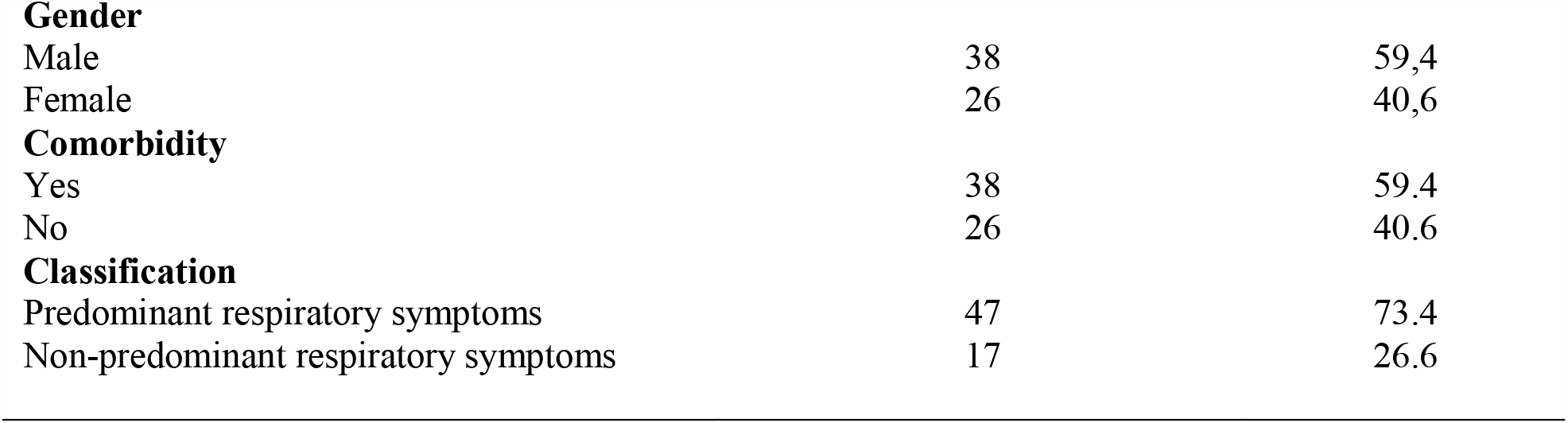
Demographic data of pediatric patients with confirmed COVID19, admitted in two hospitals of Rio de janeiro, city (March-November 15, 2020)

| Variables | N | % |
| --- | --- | --- |
| **Age** |  |  |
| 0-1 | 13 | 20.3 |
| 1-2 | 6 | 9.4 |
| 2-5 | 11 | 17.2 |
| 5-10 | 18 | 28.1 |
| 10-18 | 16 | 25 |
| **Gender** |  |  |
| Male | 38 | 59,4 |
| Female | 26 | 40,6 |
| **Comorbidity** |  |  |
| Yes | 38 | 59.4 |
| No | 26 | 40.6 |
| **Classification** |  |  |
| Predominant respiratory symptoms | 47 | 73.4 |
| Non-predominant respiratory symptoms | 17 | 26.6 |

On graph 1, we show number of confirmed cases, considering epidemiological week of beginning of the symptoms

**Graph 1.**
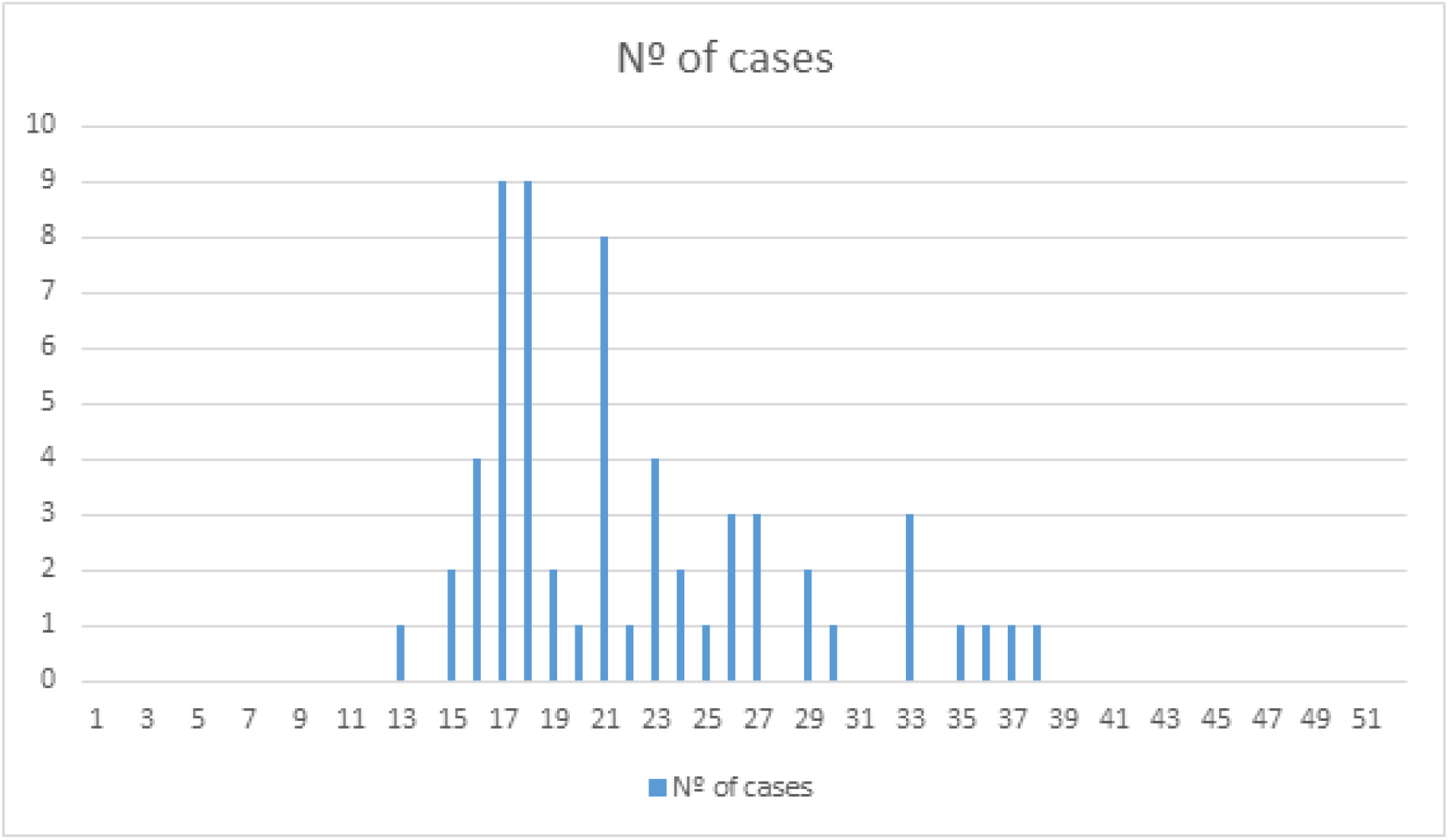
Number of pediatric confirmed cases according to the epidemiological weeks of beginning of the symptoms, in two hospitals of Rio de Janeiro city (March-November 15, 2020)

According to the main clinical manifestations, we analysed patients with predominant respiratory symptoms and patients with non-predominant respiratory symptoms. The patients were compared in relation to age, admission in intensive care units and the length of stay since the day of the beginning of the symptoms until the discharge (table 2).

**Table 2.** Comparison between pediatric patients with predominant respiratory symptoms and with non-predominant respiratory symptoms admitted in two hospitals of Rio de Janeiro, city (March-November 15, 2020)

|  | Patients<br>with<br>predominant<br>respiratory<br>symptoms | Patients<br>with<br>non-predominant<br>respiratory<br>symptoms | P value |
| --- | --- | --- | --- |

|  | N=47 | N=17 |  |
| --- | --- | --- | --- |
| Median of age (months) | 78 | 62 | 0.41 |
| Percentage of patients admitted in intensive care unit | 53,1 | 76,5 | 0.09 |
| Length of hospital stay since the beginning of symptoms (in days) | 6 | 11 | 0.08 |

In table 3, we describe the main symptoms, radiological findings, laboratory results and treatments of patients according predominant or non-predominant respiratory symptoms

**Table 3.**
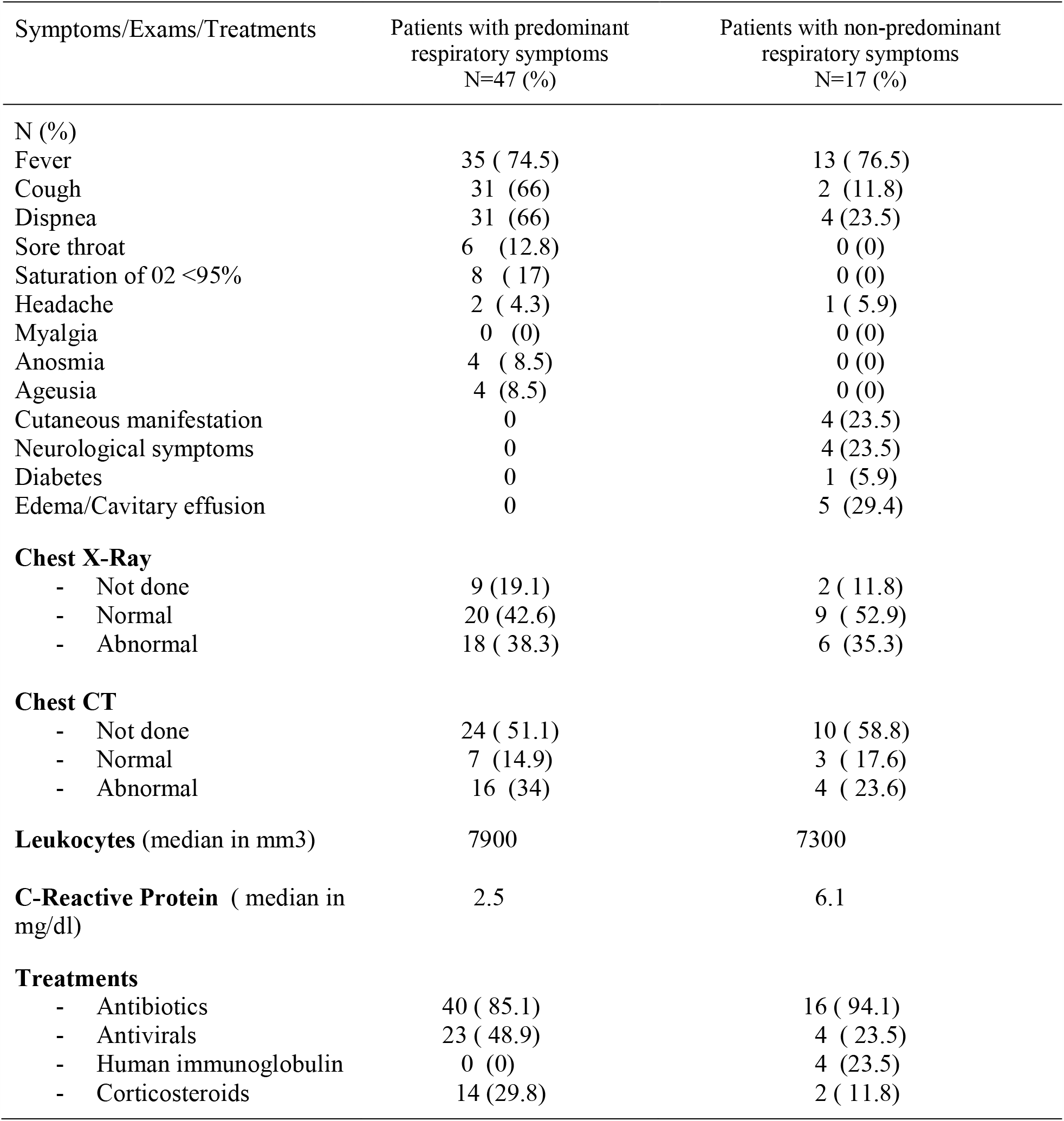
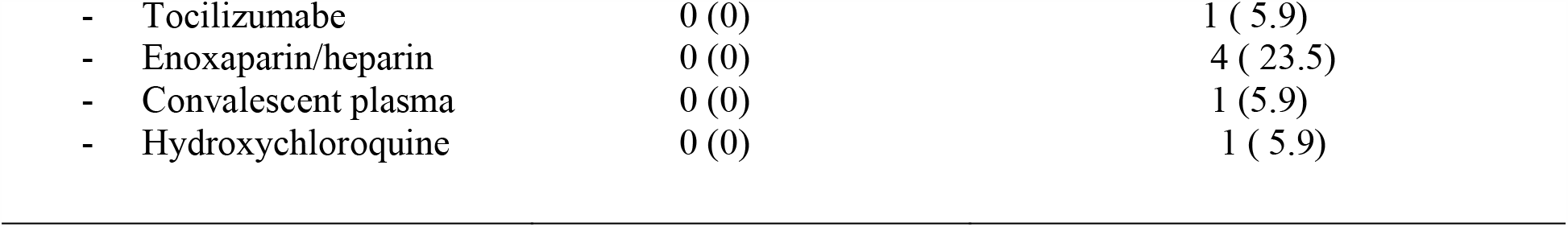
Main symptoms, laboratory results, radiological findings and treatment in pediatric patients admitted in two hospitals (March-November 15, 2020)

| Symptoms/Exams/Treatments | Patients with predominant respiratory symptoms<br>N=47 (%) | Patients with non-predominant respiratory symptoms<br>N=17 (%) |
| --- | --- | --- |
| **N (%)** |  |  |
| Fever | 35 ( 74.5) | 13 ( 76.5) |
| Cough | 31 (66) | 2 (11.8) |
| Dispnea | 31 (66) | 4 (23.5) |
| Sore throat | 6 (12.8) | 0 (0) |
| Saturation of O<sub>2</sub> <95% | 8 ( 17) | 0 (0) |
| Headache | 2 ( 4.3) | 1 ( 5.9) |
| Myalgia | 0 (0) | 0 (0) |
| Anosmia | 4 ( 8.5) | 0 (0) |
| Ageusia | 4 (8.5) | 0 (0) |
| Cutaneous manifestation | 0 | 4 (23.5) |
| Neurological symptoms | 0 | 4 (23.5) |
| Diabetes | 0 | 1 (5.9) |
| Edema/Cavitary effusion | 0 | 5 (29.4) |
| **Chest X-Ray** |  |  |
| - Not done | 9 (19.1) | 2 ( 11.8) |
| - Normal | 20 (42.6) | 9 ( 52.9) |
| - Abnormal | 18 ( 38.3) | 6 (35.3) |
| **Chest CT** |  |  |
| - Not done | 24 ( 51.1) | 10 ( 58.8) |
| - Normal | 7 (14.9) | 3 ( 17.6) |
| - Abnormal | 16 (34) | 4 ( 23.6) |
| **Leukocytes** (median in mm<sup>3</sup>) | 7900 | 7300 |
| **C-Reactive Protein** ( median in mg/dl) | 2.5 | 6.1 |
| **Treatments** |  |  |
| - Antibiotics | 40 ( 85.1) | 16 ( 94.1) |
| - Antivirals | 23 ( 48.9) | 4 ( 23.5) |
| - Human immunoglobulin | 0 (0) | 4 (23.5) |
| - Corticosteroids | 14 (29.8) | 2 ( 11.8) |
| - Tocilizumabe | 0 (0) | 1 ( 5.9) |
| - Enoxaparin/heparin | 0 (0) | 4 ( 23.5) |
| - Convalescent plasma | 0 (0) | 1 (5.9) |
| - Hydroxychloroquine | 0 (0) | 1 ( 5.9) |

The main abnormalities founded in chest radiology were: infiltrates in 12/18 (66.7%) patients and opacities in 4/18 (22.2%); and the main findings in computed tomography of chest were ground-glass opacities in 9/16 (56.2%) children, opacities in 6/16 (37.5%) and pleural effusion in 1/16 (6.3%).

The p values of leukocytes and C-Reactive Protein didn’t show difference between both groups (0.79 and 0.48), respectively.

We also analysed presence of comorbidity in patients admitted with respiratory and non-predominant respiratory symptoms and influence of comorbidity in length of stay (table 4 and 5)

**Table 5.** Presence of comorbidity in paediatric patients admitted in two hospitals of Rio de Janeiro, city (March-November 15, 2020)

|  | Yes | No | p value* |
| --- | --- | --- | --- |
| Patients with predominant respiratory symptoms | 30 | 17 | 0,22 |
| Patients with non predominant respiratory symptoms | 8 | 9 |  |

\*Chi-Square test

**Table 6.** Presence of comorbidity and length of stay more than 7 days in pediatric patients with COVID 19, admitted in two hospitals of Rio de Janeiro, city (March-November 15, 2020)

|  | Yes | No | p value * |
| --- | --- | --- | --- |
| Length of stay until 7 days | 27 | 22 | 0,20 |
| Length of stay more than 7 days | 11 | 4 |  |

\*Chi-Square test

## Discussion

After almost one year of COVID19, the world experienced difficulty to contain the spread of disease and cases are still in progress in most countries, most of them in children. Considering that Brazil is one of top leader countries in cases and deaths, despite children being less affected, knowledge about clinical manifestations, laboratory and radiological findings as well treatments are necessary in order to better understand and improve management of this population.

In our series, cases were equally distributed among different age groups of children with a median age of 5,5 years. It’s important to note that during the whole study period, schools remained closed in Rio de Janeiro city. In terms of age distribution of COVID19 positive pediatric patients, studies evaluated children in different settings. For example, Kanthimathinathan and cols evaluated children admitted in a single hospital of UK, with severe acute respiratory syndrome founding a median age of 3,5 years, while Rahba et al, studying 115 children attending in emergency room and admitted in a single hospital of São Paulo, Brazil found a median age of 2 years ^15,18^.

The cases described in this article reflected the first wave of COVID 19 in Rio de Janeiro, city where most patients admitted benning the symptoms between epidemiological week 17 and 21 (April 14 and May 23, 2020).

Due to the broad spectrum of symptoms in children studied, we analysed patients in two different categories: with predominant and non-predominant respiratory symptoms at admission. The subgroup of non-predominant respiratory symptoms represented more than one quarter of admitted patients that was compatible with a case series described by Rahba in Brazil that found 34% of gastrointestinal symptoms, but higher than reported by Zheng et al in 10 hospitals of Hubei, China where prevalence of diarreia was 12% and by Korkmaz in Turkey where vomiting and diarrhea were present in 7 % of admitted patients ^15,19,20^.

Considering patients with predominant respiratory symptoms the triad of fever, cough and dyspnea were present in more than two third of all patients. Interestingly in this group, abnormal radiological findings were present in less than 40% of patients and in many of them chest ray was not necessary, considering the favorable outcome of these patients. Computed tomography of chest was performed in less than half of patients, but the main findings as ground-glass opacities and infiltrates were similar to a large review conducted by Kumar and cols ^21^.

Inflammatory markers are usually described as normal or elevated in infection due to SARS-COV-2, depending on the ward of admission. In patients with non-multisystemic inflammatory syndrome in children (MIS-C) admitted in pediatric intensive care units in Brazil, median of leukocytes (per 1000/L) was 11,850 while in patients admitted with MIS-C was 18,275 ^12^. The values found in Spanish children also admitted in PICU was 9260 ^22^. Considering that our cases included patients admitted both in wards as in PICUs the values found were lower than reported by these authors. C-reactive protein with value > 10 mg/L was described in 68% of children admitted in a UK pediatric hospital while the median of CRP in Turkey patients were 2 mg/L^18,20^. Our median of CRP was also elevated, mainly in patients with non-predominant respiratory symptoms but without statistical significance when compared with patients with predominant respiratory symptoms.

One of the great concerns about pediatric patients is to recognize if comorbidity could influence different clinical presentations at admission or increase the length of stay. In our study, comorbidity was not a variable that influenced in favour of respiratory or non-predominant respiratory symptoms as well in length of stay higher than 7 days. Although the study was conducted in two pediatric centers it’s possible that our findings could be confirmed in a large series of cases from different institutions.

Until November 2020, there was no specific antiviral approved for COVID treatment in children patients, but some drugs such as Remdesevir or Lopinavir/Ritonavir are employed in specific settings ^23, 24^. Despite there is no gold standard for COVID 19 treatment in children, overall treatment improved since the beginning of pandemics being some therapies incorporated to therapeutics mainly for critical ill patients in pediatric intensive care units with MIS-C ^25^. In our series almost all patients received antibiotics at admission probably due to difficulty of differential diagnosis with bacterial infections. The only antiviral used was oseltamivir that was indicated in cases of acute respiratory distress syndrome due to influenza virus suspicion.

Another important question about management of our patients, was the few availability of RT-PCR tests and the high amount of them to be processed in the laboratory, during the initial months of pandemic. But even with this limitation all admitted patients were tested to COVID-19 with RT-PCR or serology in cases of patients with more than 10 days of symptoms.

Despite the study being conducted in two large pediatric hospitals of Rio de Janeiro city that could be considered a limitation, results could represent a representative panel of metropolitan areas of state, due to many patients being referred to admission from periferic hospitals. Another limitation was evaluation of only admitted patients that excluded children managed in ambulatory settings with mild disease.

## Conclusion

Non-predominant respiratory symptoms represented more than one quarter of admitted pediatric patients admitted. Comorbidity was common equally found in patients with predominant or non-predominant respiratory symptoms and his presence was not associated with a higher length of stay. Fever was the main symptom detected, elevated CRP was present and antibiotics were commonly prescribed.

## Data Availability

I declare that all data will be avaible for anaylsis

